# Mapping resources available for early identification and recovery-oriented intervention for people with psychosis in Addis Ababa, Ethiopia

**DOI:** 10.1101/2024.01.16.24301385

**Authors:** Mekonnen Tsehay, Teshome Shibre Kelkile, Wubalem Fekadu, Alex Cohen, Eleni Misganaw, Charlotte Hanlon

**Affiliations:** Department of Psychiatry and WHO Collaborating Centre for Mental Health Research and Capacity-Building, School of Medicine, College of Health Sciences, Addis Ababa University, Addis Ababa, Ethiopia; Horizon Network Zone 3, NB, Canada; Department of Psychiatry, Dalhousie University, NS, Canada; Department of Population Health, London School of Hygiene & Tropical Medicine, London, UK; Mental Health Service Users Association, Addis Ababa Ethiopia; Centre for Global Mental Health, Health Service and Population Research Department, and WHO Collaborating Centre for Mental Health Research and Training, Institute of Psychiatry, Psychology, and Neuroscience, King’s College London, London, UK; Centre for Innovative Drug Development and Therapeutic Trials for Africa, College of Health Sciences, Addis Ababa University, Addis Ababa, Ethiopia

**Keywords:** Community mental health, psychosis, traditional and faith healers, social inclusion, medical pluralism, strengths-based

## Abstract

**Background:** There is a pressing need to reduce the long duration of untreated illness and improve care and outcomes for people with psychosis in Ethiopia. This study aimed to map community resources that have the potential to be leveraged to achieve earlier and more recovery-oriented interventions for people with psychosis in Addis Ababa, Ethiopia.

**Method:** A strength-based resource mapping exercise was undertaken in two sub-cities, covering an estimated population of half a million people. We identified the types of resources to be mapped, based on their importance for multi-sectoral care in mental health: healthcare facilities, religious organisations, traditional and faith healers, non-governmental organisations (NGOs), and social/community organisations. The lead investigator traversed the study sites to gather information on community resources, recorded the Global Positioning System (GPS) coordinates of the resources, and consulted with key informants. The information obtained was complemented by a participatory Theory of Change workshop attended by 30 stakeholders.

**Results:** We identified 124 health facilities, of which only 16 health centres and nine hospitals currently provide mental health services. We identified three registered traditional healers, 38 religious organisations, 104 non-governmental organisations, and other charitable/community-based organisations. In addition, three health facilities, six holy water religious healing sites, and four traditional healers were identified as out-of-site resources that were popular and frequently visited by people living in the sub-cities. The two sub-cities also had six feeding centres each providing meals for 1000 people in need. There were extensive networks of social organisations and community-based associations. Existing care pathways are complex but commonly include traditional and religious healing sites as places of first contact.

**Conclusions:** We identified important available resources that provide a wealth of opportunities for improving the early identification and outcomes of people with psychosis.

## Introduction

In low- and middle-income countries (LMICs), the majority of people with psychosis either do not access any care or experience substantial delays in receiving care (1). An estimated 70%-90% of people with psychosis in need of mental health care in LMICs do not receive it (2, 3). According to community-based studies conducted in Ethiopia, the median duration of untreated illness in people with psychosis was more than five to seven years (4, 5). Even when care is accessed, it is often narrowly biomedical and fails to address prominent social and economic needs (6, 7).

The consequences of absent, delayed, or inadequate care for people with psychosis are substantial in terms of impairing a person’s capacity to function in social, familial, and economic domains. Furthermore, stigma, prejudice, and violations of human rights are commonly experienced by persons with psychosis, exacerbated by a lack of effective treatment (6, 8–10).

Although there are multiple factors contributing to treatment delays and the long duration of untreated psychosis, the most frequently cited relate to the demand side (e.g., financial constraints, stigma, differing explanatory models of illness) and supply-side factors (e.g. lack of available, accessible and adequate mental health services) (11, 12). Consequently, help-seeking pathways are often convoluted and may involve multiple contacts with traditional and/or faith healers or non-specialist healthcare providers before accessing mental health services (13, 14). Even when mental health services are accessed, engagement is often intermittent and poorly coordinated with pluralistic approaches to care (15, 16). There is evidence that earlier and better interventions for people with psychosis lead to better health and social outcomes, as well as lower costs (17–19).

The World Health Organization (WHO) has published guidance on person-centered and rights-based approaches to community mental health care, recommending the inclusion of communities to support pluralism of care and the provision of psychosocial interventions to augment biomedical care (20). This community-focused approach first requires an understanding of the potential contributions of both formal biomedical care and informal community-based traditional and religious healers, as well as organisations supporting social and economic well-being, to build on existing resources when designing interventions. In a previous study from rural Ethiopia, strengths-based mapping in a rural district identified rich community resources (21) that formed an input into the design of models of care for people with psychosis (22, 23). In the Ethiopia PRIME study, inter-linked interventions at the level of community, facility, and health system led to improved functioning, better socio-economic status, and reduced experiences of discrimination and abuse (4, 24). In the Ethiopia RISE trial, community-based rehabilitation led to improved functioning in people with psychosis who had not responded to primary healthcare-based mental health care (25).

Evaluation of efforts to increase access to mental health care in Ethiopia has focused on rural settings. There may be distinct opportunities and challenges in the capital city of Addis Ababa; for example, in terms of greater availability and accessibility of mental health services but also the potential for fragmentation of care, more transient populations, less social capital, and a visible population of people who are homeless and have apparent psychosis (26). Little is known about community resources in Addis Ababa and how they might be harnessed. Therefore, this study aimed to map resources that have the potential to be leveraged to achieve earlier and more holistic interventions for people with psychosis in Addis Ababa, Ethiopia.

## Methods

### Study design

We conducted a strength-based resource mapping exercise from 13^th^ September to 10^th^ October 2022. This approach includes exploring, describing, and mapping community resources before employing these resources to develop solutions to a specific problem (27). Underpinning this method is a strengths-based orientation that seeks to build on the strengths of individuals, families, and communities (28).

The study was nested within the formative phase of a larger project, SCOPE (Studying the Contexts of Recent Onset Psychoses in Ethiopia) (29). SCOPE aims to produce contextual evidence about psychosis and uses participatory methods to design and evaluate innovations to achieve earlier detection and care to improve the lives of people living with psychosis in rural and urban settings in Ethiopia.

### Setting

The urban sites included in SCOPE are Lideta and Kirkos sub-cities of Addis Ababa, the capital city of Ethiopia. According to the Central Statistical Agency of Ethiopia (CSA), in 2022 (30), the projected population of the sub-cities is 595, 973 (284, 208 in Lideta and 311, 765 in Kirkos). The total area of the sub-cities is 52,284.1 square kilometres.

### Procedures

We identified the types of resources to be mapped based on their importance for mental health: healthcare facilities, religious organisations, traditional and faith healers, non-governmental organisations (NGOs), and social/community organisations. The lead investigator then traversed the study sites to gather information on community resources, recorded the global positioning system (GPS) coordinates of the place or organization, and consulted with key informants to discover where people sought help for mental health conditions and the potential role of their organization concerning early identification and care.

We then developed an initial list of resources based on: (1) community or organization consultation; and (2) official lists of registered traditional healers and health facilities in each sub-city. Data were recorded about the total catchment population, mental health service provision (including the number of psychiatrists, mid-level mental health professionals, and psychologists), and number of general health workers who had been trained with the World Health Organization’s mental health Gap Action Programme (mhGAP) in line with the National Mental Health Strategy of Ethiopia (31). We also recorded the presence of international or national charitable organisations and social organisations (*Edir* (burial groups, supporting saving) by searching through the districts of sub-cities.

The mapping information was complemented by a participatory Theory of Change (TOC) workshop attended by 30 stakeholders. The participants were people with lived experience of mental health conditions, family members, religious leaders, traditional healers, health administration officials, community-based health workers, police, social affairs officials, NGO representatives, and community leaders. The TOC workshop aimed to obtain further information on pathways to care and existing opportunities for intervention to support people with psychosis to achieve valued outcomes. The workshop was moderated by two facilitators. Minutes were taken and used to inform the current analysis.

### Data management and analysis

Data were stored in Excel. We summarized the data descriptively (frequency and percentage) and plotted the location of the organization and facilities on maps to visualize the distribution of resources.

### Ethical considerations

We received ethical approval from the Institutional Review Board of the College of Health Science, Addis Ababa University (084/11/PSY) and Kings College London as part of a baseline situation appraisal. We also obtained permission letters from each sub-city.

## Results

Within each sub-city, we identified a wide array of community resources. These included health facilities, traditional healers, religious organisations, NGOs, and other charitable/community-based organisations. Alongside this, we identified out-of-site resources that were popular and frequently visited by people living in the sub-cities. The two sub-cities also had feeding centres, and social organisations: *Edir* (burial groups, supporting saving), *Equb* (small financial group), *Tsiwa Mahiber* (mostly religious groups), youth associations, women’s associations, elderly associations, and disability associations. Existing care pathways are complex but commonly include traditional and religious healing sites as places for both first contact and continuing contacts utilized alongside mental health services.

### Health sector

In Kirkos sub-city, there are eight public health centres, 59 private clinics, and five hospitals (three private, and two public). In Lideta sub-city, there are eight public health centres, 44 private clinics, and eight hospitals (five private, one public, the Federal police general hospital, and the Defense Forces general hospital). All public health centres and hospitals provide mental health services, except one health center in Kirkos.

### Primary mental health care

In the health centres, mental health care is provided in outpatient clinics by psychiatric nurses and masters-level mental health practitioners working alongside WHO mhGAP-trained general health care workers (nurses or health officers). Commonly the mhGAP-trained nurse or health officers were assigned to antiretroviral therapy (ART) and cancer clinic out-patient clinics. They consult mental health professionals when they see new people presenting with mental health conditions (Table 1). The patient flow is not more than 15 per month in each health centre for all mental health conditions combined, for a population served of around 35,000 people. In some health centres, mental health professionals participate in non-mental health managerial duties of the health centres rather than direct delivery of mental health care.

**Table 1:** Mental health service in health facilities of Kirkos and Lideta sub-cities.

| Health facilities in sub-cities |  | District | Catchment populations | Psychiatric Nurse | MSc Mental health practitioners | Psychiatrist | Psychologist | mhGAP trained personnel |
| --- | --- | --- | --- | --- | --- | --- | --- | --- |
| Health centres | HC-1 | KSC-D 2 | 34400 | 1 | 0 | 0 | 0 | 2 |
|  | HC-2 | KSC-D 3 | 18389 | 1 | 0 | 0 | 0 | 3 |
|  | HC-3 | KSC-D 4 | 40,000 | 1 | 0 | 0 | 0 | 4 |
|  | HC-4 | KSC-D 5 | 28999 | 1 | 0 | 0 | 0 | 1 |
|  | HC-5 | KSC-D 6 | 16771 |  |  |  |  | 1 |
|  | HC-6 | KSC-D 8 | 29430 | 0 | 0 | 0 | 0 | 0 |
|  | HC-7 | KSC-D 9 | 28914 | 1 | 0 | 0 | 0 | 2 |
|  | HC-8 | KSC-D 11 | 30108 | 1 | 0 | 0 | 0 | 0 |
|  | HC-9 | LSC-D 1 | 25922 | 1 | 0 | 0 | 0 | 3 |
|  | HC-10 | LSC-D 7 | 97,617 | 0 | 2 | 0 | 0 | 10 |
|  | HC-11 | LSC-D 3 | 51,000 | 2 | 0 | 0 | 0 | 3 |
|  | HC-12 | LSC-D 2 | 25684 | 1 | 0 | 0 | 0 | 5 |
|  | HC-13 | LSC-D 9 | 31113 | 1 | 0 | 0 | 0 | 4 |
|  | HC-14 | LSC-D 10 | 39763 | 2 | 0 | 0 | 0 | 3 |
|  | HC-15 | LSC-D 8 | 48,000 | 1 | 0 | 0 | 0 | 3 |
|  | HC-16 | LSC-D 4 | 25,817 | 1 | 0 | 0 | 0 | 0 |
| Public hospital | GH-1 | KSC-D 7 | - | 7 | 0 | 2 | 1 | 3 |
|  | GH-3 | LSC-D 7 | - | 0 | 0 | 1 | 0 | 0 |
|  | GH-4 | LSC-D 7 | - | 5 | 0 | 23 | 7 | 0 |
| Private hospitals | PH-1 | KSC-D 7 | - | 0 | 0 | 1 | 0 | 0 |
|  | PH-2 | KSC-D 9 | - | 0 | 0 | 2 | 0 | 0 |
|  | PH-3 | KSC-D 4 | - | 0 | 0 | 2 | 0 | 0 |
|  | PH-4 | LSC-D 10 | - | 0 | 0 | 1 | 0 | 0 |
|  | PH-5 | LSC-D 3 | - | 0 | 0 | 2 | 0 | 0 |
|  | PH-6 | LSC-D 4 | - | 0 | 0 | 2 | 0 | 0 |
|  | PH-7 | LSC-D 7 | - | 1 | 0 | 0 | 1 | 0 |

HC=health center, GH=Government/public hospital, PH=private hospital, mhGAP=mental health gap action programme, LSC-D=Lideta sub-city district, KSC-D=Kirkos Sub-city district

### Hospital-level secondary and tertiary mental health services

Black Lion Comprehensive Specialized Hospital (Lideta sub-city) has an outpatient psychiatric clinic staffed with 23 psychiatrists, five psychiatric nurses, and seven clinical psychologists. Zewditu Memorial Referral Hospital (Kirkos sub-city) has an outpatient psychiatric unit, two crisis admission beds, and a four-bed unit for people with substance use disorders. Psychotherapy services are also provided in these hospitals.

In private hospitals, outpatient and in-patient mental health services are provided by part-time psychiatrists. The Federal Police Hospital and the Ethiopian Defense Force Hospitals have psychiatric services but currently, the service is only for the military.

Although outside the two sub-cities, referrals are made to Amanuel Mental Health specialized hospital, a dedicated public mental health hospital with 239 beds, 27 emergency beds, an in-patient substance use unit, and multiple outpatient clinics with 128, 000 people with MHCs seen per year. The two main private psychiatric clinics in Addis Ababa are also outside the sub-cities but accessed by inhabitants (Figure 1).

**Figure 1.**
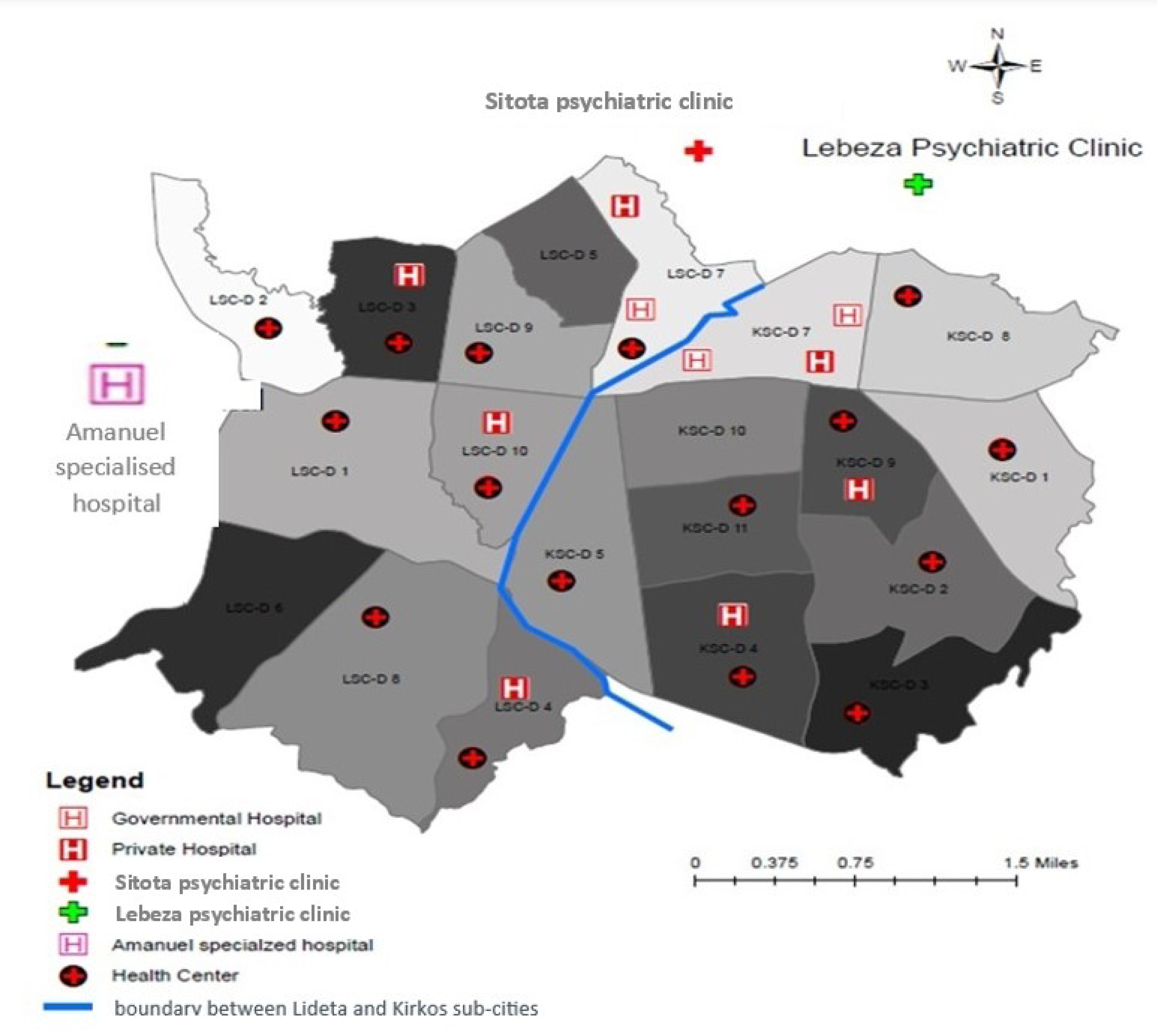
**Health care facilities within the study area and popular facilities outside the study area**: LSC-D=Lideta sub-city districts, KSC-D=Kirkos sub-city districts

### Religious, traditional, and faith healer (TFH) sites

Within the two sub-cities, we identified three traditional healers (mostly herbalists but also practicing spiritual or faith-based healing, diviner, giving counsel, and bone setting) and four holy water healing sites linked to the Ethiopian Orthodox Church. Holy water is typically the first port of call when a family member has a condition perceived to be triggered by a spiritual or supernatural problem, an attribution which is commonly linked to severe mental illness. People drink the holy water or are baptised. They may stay at the holy water site for extended periods where they are supported by family or overseen by a holy water attendant who is paid for the role (Table 2).

**Table 2:** Religious organisations and their mental health care provision.

| Religious organisations | Religious healing practices |
| --- | --- |
| Ethiopian Orthodox Christian churches<br>Eleven churches with/without holy water<br>Four holy water healing sites | Pray and recite verses from the holy bible for the ill.<br>Bless the water and administer it as holy water.<br>Some have holy water sites visited by several people every day. E.g.: Tekle haymanot, Yohanis, Lideta and St. Mary, and St. Egziabher Ab holy water sites. |
| Nine Muslim mosques | The sheik recites verses from the holy Quran for the ill.<br>Outside of our study area, at the Quran home, the holy Quran will be recited by a gifted person |
| Thirteen Protestant Christian churches | Advertise at least one day per week for faith healing programs. |
| Two Catholic churches | Priests, recite verses from the holy bible and give “kibakdus”, holy anointing for the ill. |
| Three Traditional healers | Traditional healers are often consulted by people with mental illness. They are mostly herbalists, but they practice also spiritual or faith-based healing, divining, advice on what to do, and bone setting) |

There are also other popular holy water places outside the two sub-cities. People also use the most popular Muslim religious healing site outside of our study area, the Quran home. The holy Quran will be recited by a person recognized as having gifts of healing. There are 11 Ethiopian Orthodox Christian churches, 13 protestant churches, nine mosques, and two catholic churches in the sub-cities (Figure 2).

**Figure 2.**
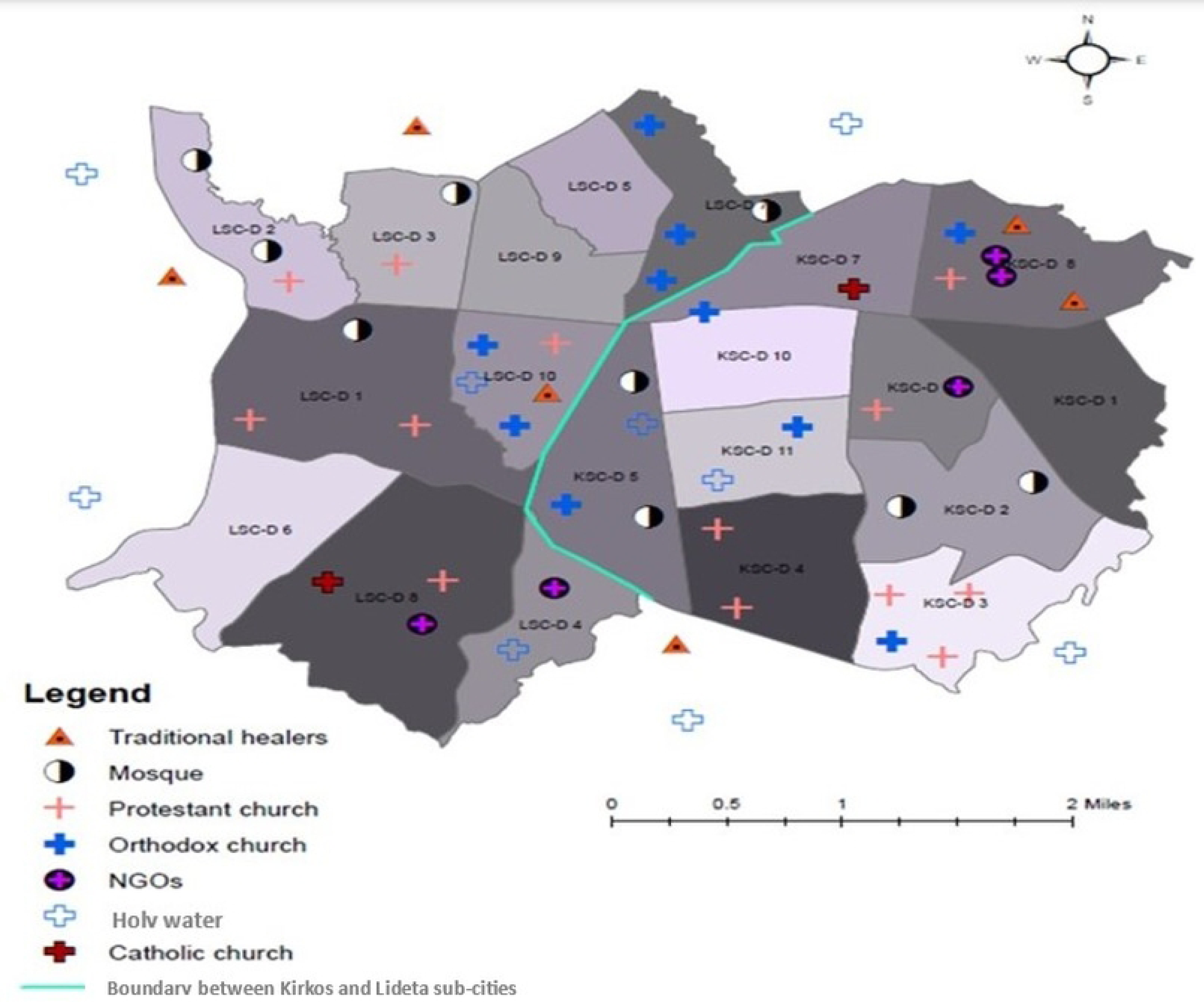
**Religious organisations, non-governmental organisations, and traditional and faith healers’ sites within and outside the study area**: LSC-D=Lideta sub-city districts, KSC-D=Kirkos sub-city districts

### Non-governmental and community-based organisations

There are more than 104 charitable organisations registered in the two sub-cities. Makedonia, Selihom, and Gergesenon charitable organisations provide social care and residential services to people with mental health conditions whose families cannot cope or who have no social support. They employ master’s and degree-level mental health care providers and also work in collaboration with local mental health services for case reviews by senior psychiatrists.

### Community/social organisations

Numerous community-based organisations exist. The first is *Edir*, mainly a funeral association that provides financial, practical, and emotional support when a family member dies. However, the role of *Edir* groups also includes the provision of support to members experiencing various social, economic, and health problems. There are an estimated 51 *Edir* associations in each district in the Lideta and Kirkos sub-cities. Each group has about 100–620 households as members. Individuals can be members of more than one *Edir* association.

*Tsiwa Mahber* is the second most common type of organization, specific to the Orthodox Christian church. Activities include group prayer, singing, and shared meals (called “Tsebel tsediq” in Amharic) taking place once per month in a member’s home or church compound. The name of a specific *Tsiwa Mahber* association is usually taken from a saint’s name. The number of members may range from 10 to 30.

In each of the two sub-cities, there are also many other associations, each with a specific focus: for young people, women, the elderly, and people with disabilities. Government representatives directly or indirectly coordinate these non-profit community-based associations. Associations for the elderly and those with disabilities aim to provide mutual support for members, provide them with more authority, and uphold their legal rights in society. Members will be the first to receive funding when it becomes available from relief organisations or the government. Women’s associations work to advance women’s rights, increase their participation in family and community decision-making, and strengthen their economic position. The purpose of the youth associations is to promote participation in neighborhood events, access to skill development opportunities, and projects that generate income.

### Feeding centres

The Addis Ababa city administration has established feeding centres throughout the city, to feed those who are unable to obtain a meal at least once per day. The program is coordinated by the city administration and funded by volunteers (Table 3).

**Table 3:**
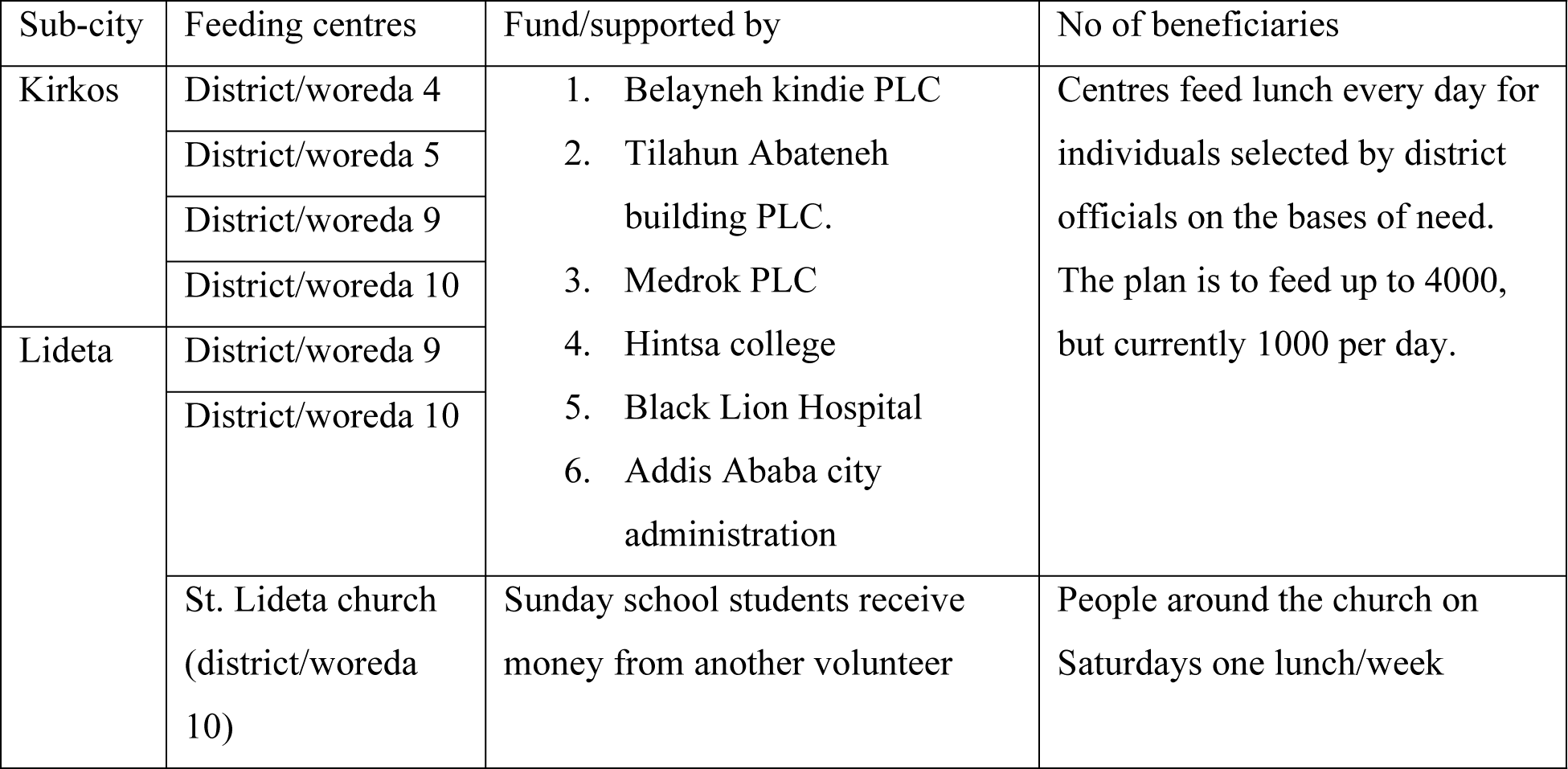
Feeding centres in the SCOPE study area (Kirkos and Lideta sub-cities of Addis Ababa)

We found that certain community-based organisations and associations are not commonly accessible for people with severe mental illness for example, Edir, Equb, youth associations, women’s associations, elderly associations, and disability associations. With respect to non-governmental and charitable organisations, target populations for the organization are usually circumscribed (e.g., for women and children) and often do not include people with severe mental illness. In feeding centres, people with severe mental illness were able to obtain meal cards from district officials, as long as they were not acutely unwell. Others could also request/apply for meal cards for them on their behalf.

### Theory of Change workshop

The initial sources of assistance for people with severe mental illness were identified as community resources, primarily religious healers, holy water sites, and a new outreach service from primary health care facilities (“family health teams”). To help with community-based case identification, access to care, and to support holistic recovery for a person with psychosis, collaboration and the establishment of referral systems were recommended.

## Discussion

In this study, we aimed to map available mental health care resources in two sub-cities of Addis Ababa to inform improved care planning for people with psychosis. We identified health facilities, registered traditional healers, religious organisations, and charitable/community-based organisations. We also found popular resources outside of the two sub-cities. Feeding centres, social organisations youth associations, women’s associations, elderly associations, and disability associations were also considered important but not always available to people with psychosis and their families. Traditional and faith healing sites are commonly the places of first contact.

Similar to a resource mapping exercise conducted in a rural Ethiopian district, there are rich cultural and social resources that may contribute to the recovery of people with psychosis in Addis Ababa (32). Our findings of many diverse community organisations indicated that ties to local community structures remain salient in this capital city, contrary to our expectations.

Nonetheless, many community supports appear not to be fully accessible to people with severe mental illness. More in-depth research is required to understand the reasons for this. In rural areas, there is evidence that stigma and impoverishment fuel the exclusion of people with psychosis from community life (33, 34). Even disability organisations that would be expected to address the needs of people with psychosis define themselves narrowly as only catering to the needs of people with physical disabilities. A fruitful strategy may, therefore, focus on stigma reduction alongside advocacy directed at organizations and services that have the most potential to provide services for people with MHCs and their families. The recent Lancet Commission on ending mental health stigma and discrimination (35), identified social contact interventions involving people with lived experience of MHCs to be the most effective way to reduce stigma.

In the SCOPE project, within which this study is nested, we are working with the Mental Health Service User Association of Ethiopia to develop such interventions to overcome exclusion.

Religious and traditional healing are integral aspects of care for many people with psychosis in Ethiopia. Our context mapping identified numerous and important healing sites within and around Addis Ababa where people with psychosis and their families seek help. Holy water sites linked to the Ethiopian Orthodox Church, charismatic Protestant churches, and Mosques all provided faith healing. Alongside religious approaches were a variety of traditional healers using herbs, medicines, and eclectic approaches to healing. Although mental health services in Addis Ababa are much more accessible than in rural areas, religious and traditional approaches are usually the first port of call. In an old study conducted at the main psychiatric hospital, Amanuel Specialized Mental Hospital, only 41% of service users came directly to the hospital, with the remaining trying alternatives first (13). In Ethiopia, people with psychosis and their families commonly seek out biomedical mental healthcare services after investing a substantial amount of time and money in alternative places (13, 14, 36). This concern was also raised by the stakeholders in our Theory of Change workshop. However, compared to rural areas, little is known about the experience and role of community-based traditional and faith-healing providers and how people with psychosis and their families navigate help-seeking journeys in Addis Ababa.

Our mapping study also indicated that efforts to decentralize mental health care through integration in primary health care facilities have not yet achieved their potential in Addis Ababa.

At present the patient flow for mental health care in primary health care centres is very low relative to projected population needs, while the specialist mental health services are overwhelmed. There are many potential advantages to the integration of mental health in primary health care services: accessibility, efficiency, affordability, acceptability, and greater potential for care that attends to both physical and mental health needs. Previous studies from Ethiopia have shown high levels of premature mortality in people with psychosis which is mostly caused by poor physical health (37).

A study from Ethiopia examining the impact of the COVID-19 pandemic on mental health services concluded that more integrated, de-centralized mental health care would have made the mental health system more resilient and should be urgently prioritized(38).

In rural settings in Ethiopia, task-shared mental health care within primary health care for people with psychosis has been shown to be safe and effective (39) but sustainability is limited by system constraints, including staff turnover, medication supply interruptions, and the lack of ongoing support from secondary mental health care services (40). In Addis Ababa, these system bottlenecks are less salient, but fragmentation of services is evident and currently undermines successful de-centralization. Nonetheless, the recent Ministry of Health initiative to develop “family health teams” could play a key role in improving care for people with psychosis. Family health teams provide community outreach from primary healthcare facilities, specifically focused on those who are vulnerable and would otherwise struggle to access care, including homeless populations. Given their prominent community footing, family health teams could contribute importantly to the early identification and ongoing engagement of people with psychosis in care.

Although we followed robust methods to map the resources, there were some limitations to our study. We only included registered and well-known traditional and faith healers in the study area, but we assume that more traditional healers may exist. It was also difficult to know the number of social organisations, such as *Tsiwa Mahber* and *Equb*.

## Conclusions

We identified important community resources that may be used by people with severe mental illness. Since many people may have access to these resources, these approaches may be scalable. However, awareness-raising and stigma reduction efforts are needed to facilitate access for people with psychosis. Integrating community resources into mental health care can be a comprehensive, cost-effective, and long-lasting strategy.

## Data Availability

All relevant data are within the manuscript and its Supporting Information files.

## Author Contributions

This study was conceived by CH and MT and designed by: TS, WF, AC, EM, and CH. Data were collected and analyzed by MT. The paper was drafted by MT and reviewed by TS, WF, AC, EM, and CH. All authors reviewed the final draft and approved the submission.

## Acknowledgments

We are grateful for the support provided by the Addis Ababa sub-city administration, district administration, the health office, the health center workers, community leaders, and the community that participated as key informants. CH receives support from the National Institute for Health and Care Research (NIHR) through the NIHR Global Health Research Group on

Homelessness and Mental Health in Africa (NIHR134325) and the SPARK project (NIHR200842) using UK aid from the UK Government. The views expressed in this publication are those of the authors and not necessarily those of the NIHR or the Department of Health and Social Care. CH also receives support from WT grants 222154/Z20/Z and 223615/Z/21/Z.

## Supporting information

**S1 Fig. Protestant Christian church program adverts**

**S2 Fig. Type of herbal plants posted at the traditional healer healing site**

**S3 Fig. St Mary’s Orthodox Christian Church, Entoto, Addis Ababa, Ethiopia**

